# Critically ill patients with a reverse blood pressure dipping phenotype at increased risk for delirium and death

**DOI:** 10.1101/2025.01.28.25321270

**Authors:** Nadim El Jamal, Thomas G. Brooks, Antonijo Mrcela, Michael V. Genuardi, Garret A. FitzGerald, Carsten Skarke

## Abstract

**Background:** The ICU environment is disruptive to a patient’s biological rhythms where sleep-wake cycles are often desynchronized from the environmental day-night changes. This puts patients at increased risk to develop delirium with consequent fiscal pressure for the health care system. An underappreciated dimension is how time-specific patient phenotypes in the critical care environment relate to clinical outcomes. We set out to analyze how rhythmic components (or the lack thereof) in physiological data streams sampled at high resolution in the ICU were associated with the future incidence of delirium and death. To offer cues for further interrogation into mechanism and risk prognosis, we examined differences in 24-hour fluctuations of clinical labs in ICU patient populations at risk.

**Methods:** Rhythmic components using dipping ratios and JTK_CYCLE statistics were derived from 24-hour blood pressure and heart rate measurements available from ICU patient admissions recorded in the MIMIC IV database. Logistic adjusted regression models assessed the association between disrupted vital sign rhythms and the future incidence of delirium during the same hospital admission and death. Aggregation of numeric clinical lab measurements across the first 24 hours from all patient admissions allowed modelling of rhythmic patterns and subsequent association studies to link potential biochemical mechanisms to perturbed vital sign rhythms and adverse ICU outcomes.

**Results:** Patients with reverse blood pressure dipping were at a 40% higher risk to have a diagnosis of delirium (Odds Ratio: 1.40, 95% CI: 1.14-1.72) and a 13% increased risk of death (Odds Ratio: 1.13, 95% CI: 1.02-1.26). Compared to the patient population with nocturnal blood pressure dip, reverse dippers showed 24-hour biochemistry profiles suggestive of altered circadian programs specifically in clinical parameters of renal, metabolic, and hemostatic function.

**Conclusions:** Reverse blood pressure dipping can be an early sign for the future development of delirium in the ICU and is accompanied by disrupted biorhythms across multiple organ systems. Dampened and reversed heart rate and blood pressure rhythms are associated with a higher risk for death in ICU patients. Considering the inclusion of these risk factors in preventive care may improve patient outcomes and reduce burden on the health care system.

## Background

The environment in the intensive care unit (ICU) with frequent checkups, procedures and abundant light and sound exposure is disruptive to a patient’s biological rhythms. The most common concern in the ICU are sleep-wake cycles desynchronized from the environmental day-night changes. Large-scale quantification of disrupted sleep-wake cycles remains a challenge and requires dedicated tools ranging from surveys to wearable devices(1). However, physiological oscillators, such as heart rate, blood pressure, and body temperature, are recorded in the ICU at high frequencies(1–3) and may serve as proxy features to quantify the disruption of diurnal rhythms, allowing evaluation of the association of circadian disruption with clinical outcomes.

About a third of critically ill patients admitted to the ICU develop delirium, a condition described as a mental state of confusion with reduced awareness of surroundings.(4) Characteristic is a sudden onset over hours to days, and patients diagnosed with delirium face an increased incidence of dementia and mortality among other poor outcomes.(4–6) Delirium is multifactorial but notable risk factors for developing this condition include day to night disorientation and sleep deprivation which both indicate disrupted circadian programs.(4, 7) Longer hospital admissions, additional outpatient visits and increased needs for rehabilitation put an enormous strain on the health care system. The costs attributable to this have been estimated to amount to hundreds of billions of US dollars.(8)

Parsing diurnal variability in time series data allows to identify time-specific differences and their association to disease outcomes. In 24-hour blood pressure monitoring, for example, the nocturnal dip is a commonly used metric to gauge a patient’s risk to develop cardiovascular and kidney disease.(9–12) However, more comprehensive methods to interrogate daily oscillations have been developed. Adopting the JTK_CYCLE algorithm that assesses the significance of rhythmic components, defined as repeat cycles in time series data where a steady increase towards a peak is followed by a steady decrease towards a trough, (13) we were able to improve risk stratification for patients with chronic renal insufficiency based on their 24-hour blood pressure profile.(14) For large cohort studies, we demonstrated the utility of rhythm measurement by associating dampened amplitudes of wrist temperature traces with the future onset of 73 diseases.(15) We extended this work to ask if we can identify rhythmic components in time series data collected in the ICU environment and how these relate to clinical outcomes.

We leveraged MIMIC IV as an electronic health record dataset to parse time-specific vital signs and patient outcomes. We found that in the ICU environment, patients with reverse blood pressure dipping – known to be a high-risk phenotype for CVD – were at higher odds to be diagnosed in the future with delirium and to have higher mortality compared to patients with blood pressure dipping at night. Furthermore, we discerned diurnal fluctuations in clinical labs and found perturbed rhythms in markers of kidney metabolism and hemostasis which offer cues for further interrogation into mechanism and risk prognosis.

## Methods

### The MIMIC-IV Database

The Medical Information Mart for Intensive Care-IV (MIMIC-IV) database contains Electronic Health Record data of real ICU patient admissions from 73,181 ICU admissions at the Beth-Israel Deaconess Medical Center in Boston between 2008 and 2019.(16, 17) It contains vital signs recorded in patient charts at hourly intervals embedded in comprehensive in-hospital data such as diagnoses, procedures, diagnostic tests, medications, and intakes/outputs. Each patient is also followed up for out of hospital mortality for a maximum of 1 year post discharge from the last admission on record by tracking social security mortality data. The data were de-identified to maintain HIPAA standards with identifiers replaced by random ciphers. Dates were also shifted randomly such that a single date shift is applied to all data from the same individual. However, time of day and seasonality were preserved. We pulled the data from the study website (https://physionet.org/content/mimiciv/2.2/, version 2.2, downloaded January 5, 2023).

### Rhythmic Assessments

We extracted the hourly recordings of systolic blood pressure (both arterial and non-invasive), temperature (the body site of measurement is not further specified), and heart rate. We excluded any readings that were outside the range of 50 mmHg to 200 mmHg for systolic blood pressure, 30 °C to 43 °C for temperature, and 50 bpm to 150 bpm for heart rate. These criteria eliminated 0.25% of all blood pressure readings, 0.08% of temperature readings, and 1.17% of heart rate readings. For each individual vital sign, we only included ICU admissions that had at least 14 daytime and 6 nighttime measurements to ensure an representative coverage of both day and night.(10) We specified nighttime as the interval between midnight and 6 am and daytime as that between 6 am and midnight. Because only 20% of ICU admissions fulfilled our inclusion criteria for temperature measurements, we excluded temperature from further analysis to decrease the risk of selection bias (Figure S1 bottom panel). We calculated non-parametric Jonckheere-Terpstra-Kendall (JTK) *p*-values, amplitude, as well dipping ratio (nighttime mean divided by daytime mean) for included admissions. Values for each admission were binned by hour of the day and the mean of each bin was used as input to JTK, thereby ensuring all admissions had between 20 and 24 datapoints, allowing *p*-values comparisons across admissions.(14) We used the JTK p-value to determine rhythmicity for each time series of vital signs (p≤0.05).

To account for non-linear associations with amplitude, we categorized amplitude based on the quartiles of the studied populations (Table S1). Dipping status for both blood pressure and heart rate was categorized as dippers (dipping ratio ≤0.9), non-dippers (0.9< dipping ratio ≤1) and reverse dippers (dipping ratio >1).(10) To study whether the disruption of rhythms in the first 24 hours is associated with future adverse outcomes, we limited our study to vital sign data collected only during the first 24 hours of ICU admission. Admissions with less than 24 hours between first and last measurements were excluded from analysis. For each analysis, patients fulfilling our rhythm quality control criteria for more than one admission had one of their admissions chosen at random for inclusion.

### Clinical Outcomes and Statistical Analyses

Incident delirium was the primary outcome of interest. MIMIC-IV reports results from each component of the confusion assessment method (CAM).(18) As per CAM guidelines, we defined delirium as the presence of both a fluctuation in mental status from baseline and inattention and the presence of either a decreased level of consciousness or disorganized thinking.(18) For our delirium analysis, we included patients with a negative CAM in the first 24 hours and with a repeat CAM after the first 24 hours. We then ran a logistic regression model containing each tested rhythmic parameter individually with the components of the OASIS score (Oxford Acute Severity of Illness Score); variables: Pre-ICU length of stay, age, Glasgow coma scale, heart rate, mean arterial pressure (MAP), respiratory rate, temperature, urine output, mechanical ventilation, elective surgery.(19) To adjust better, vital sign components were computed as actual 24-hour averages and included in the model with normal cubic spline functions; other components were derived from the PostgreSQL code provided by the MIMIC code repository.(20) We also studied mortality during the ICU visit or up to one month (30 days) afterwards from the day of admission using the same methodology. Resulting *p*-values for both outcome analyses were adjusted for multiple testing by the Benjamini-Hochberg method.(21) A *q*-value cutoff of 0.1 was considered as the threshold for statistical significance.

To explore rhythms in biochemical processes and their association with vital sign rhythms we aggregated all numeric clinical lab measurements across the first 24 hours from all patients with a vital sign rhythm pattern of interest after removing outlier values (beyond 1.5 times the second and third quartiles) and labs with less than 100 samples. If a patient had multiple visits, all were included, and dipping status was determined for each visit separately. We then ran a Cosinor analysis, with random intercept effects for subjects and admissions, using the lme4 R package.(22) To test for amplitude and rhythm differences among the patient groups, we used an F-test with the lmerTest package and Kenward-Rogers degrees of freedom to compare the nested model where both groups (dipping or reverse dipping) had separate cosinor fits to the model where all groups had the same cosinor fit but separate intercepts.(23) We considered a *q*-value cutoff of 0.1 as the threshold for significant rhythm differences across compared groups.

## Results

### Patient Data Base Characteristics

Data recordings from most ICU patient admissions met the day-/nighttime coverage criteria for blood pressure (45,287 out of 73,181, 61.9%) and for heart rate (47,130 out of 73,181, 64.4%), (Figures S1 and S2). Illness severity parameters showed that a less healthy population was included and analyzed after filtering for rhythm quality control criteria for both heart rate and blood pressure (Table 1). After excluding repeat stays and stays with unmet CAM criteria, 7,346 out of 73,181 (10%) and 7,552 out of 73,181 (10.3%) patient admissions remained for blood pressure and heart rate, respectively, which also qualified for delirium analysis (Figure S2). Most patients (76%) had only one ICU admission (Figure S3). A total of 33,970 (46.4% of all admissions) and 35,101 (48% of all admissions) admissions were submitted to the mortality analysis for blood pressure (BP) and heart rate (HR), respectively (Figure S4).

**Table 1.** Patient characteristics collected during the first 24 hours for comparing admissions in the MIMIC IV database fulfilling quality control criteria of vital sign measurements to those that do not fulfill the criteria. **GCS:** Glasgow Coma Scale, **MAP:** Mean Arterial Pressure.

|  | BP Rhythm<br>Inclusion<br>Criteria<br>Absent<br>(n=27,597) | BP Rhythm<br>Inclusion<br>Criteria<br>Present<br>(n=45,287) |  | HR Rhythm<br>Inclusion<br>Criteria Absent<br>(n=25,936) | HR Rhythm<br>Inclusion Criteria<br>Present<br>(n=47,130) |  |
| --- | --- | --- | --- | --- | --- | --- |
| Parameter | Mean (SD), or<br>n (%) | Mean (SD), or<br>n (%) | *p-*<br>*Value* | Mean (SD), or n<br>(%) | Mean (SD), or n<br>(%) | *p-*<br>*Value* |
| OASIS<br>score | 28.81 (8.58) | 31.40 (8.42) | <0.001 | 28.70 (8.53) | 31.36 (8.47) | <0.001 |
| Age (years) | 63.44 (17.49) | 65.43 (16.46) | <0.001 | 63.66 (17.42) | 65.23 (16.55) | <0.001 |
| Race |  |  | <0.001 |  |  | 0.006 |
| American<br>Indian or<br>Alaska<br>Native | 46 ( 0.2) | 92 ( 0.2) |  | 42 ( 0.2) | 96 ( 0.2) |  |
| Asian | 838 ( 3.0) | 1312 ( 2.9) |  | 785 ( 3.0) | 1364 ( 2.9) |  |
| Black | 3124 (11.4) | 4795 (10.6) |  | 2884 (11.1) | 5060 (10.7) |  |
| More than<br>one race | 22 ( 0.1) | 46 ( 0.1) |  | 23 ( 0.1) | 45 ( 0.1) |  |
| Native<br>Hawaiian or<br>Other<br>Pacific<br>Islander | 48 ( 0.2) | 62 ( 0.1) |  | 44 ( 0.2) | 66 ( 0.1) |  |
| Other | 902 ( 3.3) | 1459 ( 3.2) |  | 854 ( 3.3) | 1511 ( 3.2) |  |
| Unknown | 2729 ( 9.9) | 4911 (10.8) |  | 2568 ( 9.9) | 5101 (10.8) |  |
| White | 19808 (72.0) | 32610 (72.0) |  | 18707 (72.2) | 33887 (71.9) |  |
| Sex (M) | 15178 (55.2) | 25411 (56.1) | 0.012 | 14480 (55.9) | 26253 (55.7) | 0.628 |
| Pre-ICU<br>length of<br>stay (hours) | 2147.19<br>(6893.04) | 3463.88<br>(10243.00) | <0.001 | 2151.87<br>(7048.70) | 3423.31<br>(10097.05) | <0.001 |
| (days) | 89.46 (287.2) | 144.33<br>(426.79) |  | 89.66 (293.70) | 142.64 (420.71) |  |
| GCS | 13.93 (2.41) | 13.52 (2.73) | <0.001 | 13.94 (2.41) | 13.53 (2.73) | <0.001 |
| Heart rate<br>(bpm) | 83.63 (14.97) | 85.87 (15.52) | <0.001 | 82.59 (15.54) | 86.39 (15.09) | <0.001 |
| SBP<br>(mmHg) | 119.44 (17.24) | 117.73 (16.04) | <0.001 | 119.36 (16.90) | 117.83 (16.28) | <0.001 |
| Respiratory<br>rate (/min) | 24.40 (9.38) | 26.55 (9.47) | <0.001 | 24.07 (9.41) | 26.65 (9.42) | <0.001 |
| Temperature<br>(Celsius) | 36.77 (0.51) | 36.86 (0.50) | <0.001 | 36.76 (0.52) | 36.87 (0.50) | <0.001 |
| Urine output<br>(ml) | 1553.69<br>(1146.38) | 1880.69<br>(1278.78) | <0.001 | 1531.32<br>(1113.07) | 1882.27<br>(1290.12) | <0.001 |
| Mechanical<br>ventilation<br>(%) | 6973 (25.3) | 17603 (38.9) | <0.001 | 6616 (25.5) | 18068 (38.3) | <0.001 |
| Elective<br>surgery (%) | 764 ( 2.8) | 1200 ( 2.6) | 0.318 | 747 ( 2.9) | 1226 ( 2.6) | 0.026 |

**Table 2.**
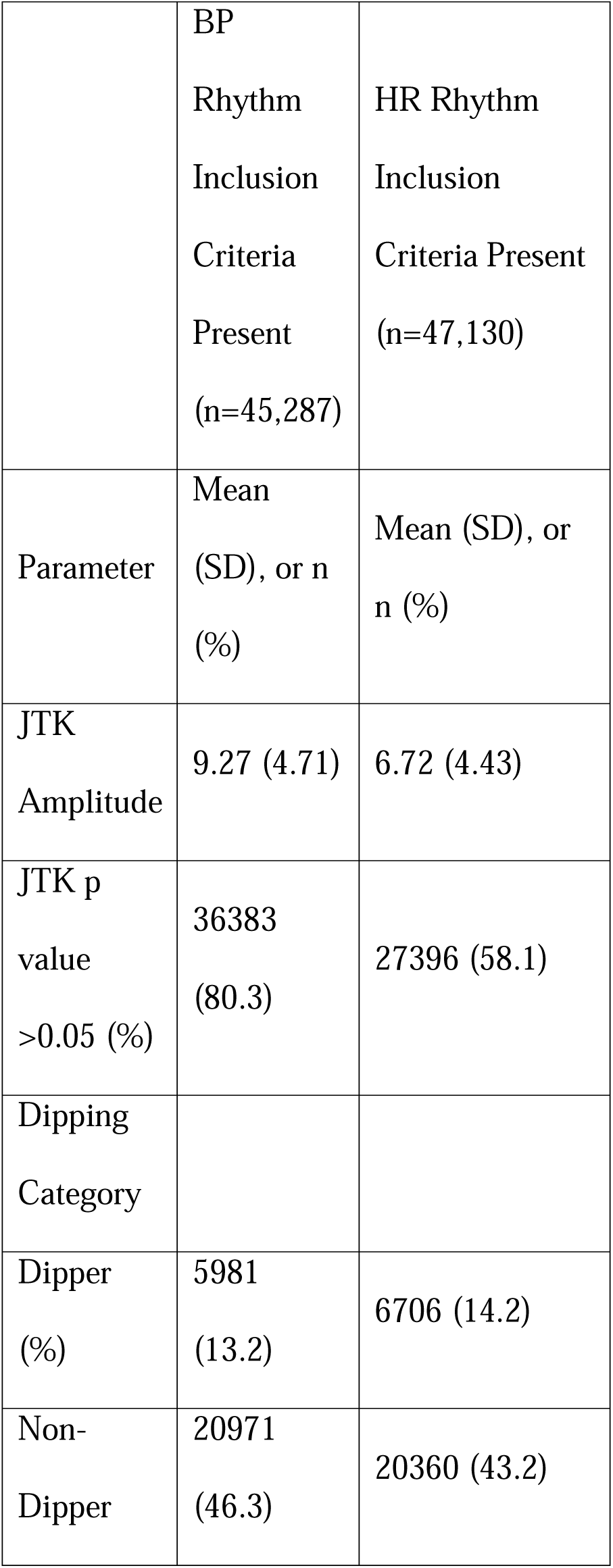
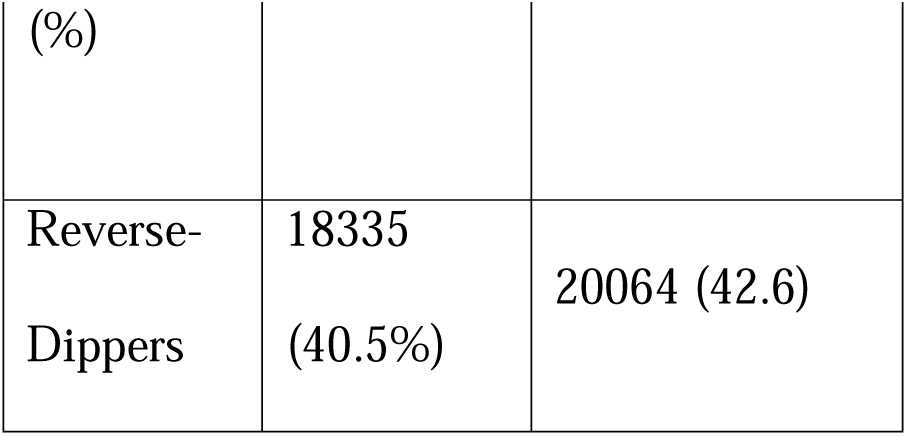
Rhythmic parameter summary statistics for admissions fulfilling rhythm quality control criteria. Rhythmic parameter summary statistics are not possible in excluded visits due to low result confidence in time series data that does not fulfil quality control criteria.

### Reverse blood pressure dipping phenotype predictive of future diagnosis of delirium

Incident delirium was diagnosed in 15.9% (1,171 out of 7,346) and 15.7% (1,189 out of 7,552) of the included BP and HR ICU admissions, respectively. (Figure S2). Patients with a reverse blood pressure dipping phenotype had a 40% greater risk for developing delirium during their ICU admission (Odds Ratio: 1.40, 95% CI: 1.14-1.72, *q*=0.017, Figure 1 left, Table S2) compared to patients with a dipping blood pressure profile. Patients with a non-dipping BP profile, in contrast, did not show this association when compared to BP dippers (Odds Ratio: 1.01, 95% CI 0.81-1.24, *q*=0.971). Assessment of rhythmic components of BP suggested a link between non-oscillating blood pressure profile (*JTK p*-value>0.05) and delirium (19% elevated risk), but this relationship has a higher false discovery rate (Odds Ratio: 1.19, 95%CI: 0.99-1.41, *q=*0.238). Though higher amplitudes of blood pressure oscillations showed a lower risk of delirium incidence compared to low amplitudes, this effect was small and non-significant (Odds Ratio: 0.94, 95%CI: 0.77-1.14, *q=*0.693 comparing JTK Amplitude Q1 versus Q4 in Figure 1 left).

**Figure 1.**
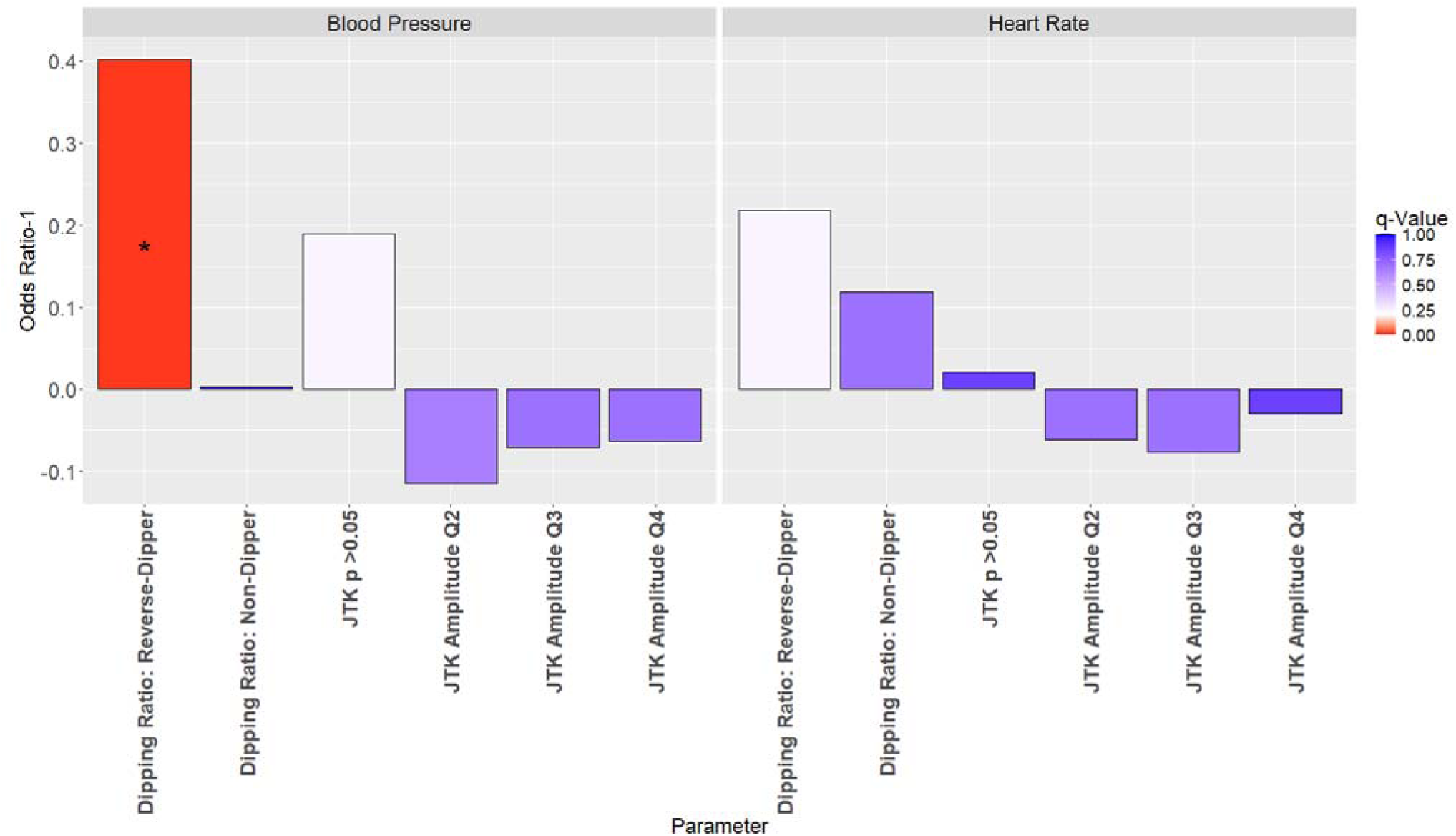
Associations between different rhythm parameters and the future development of delirium. The left panel displays results from the logistic regression models testing blood pressure rhythmic parameters, the right panel displays results from heart rate rhythmic parameters. Reference categories are Dippers, JTK *p-*value ≤0.05, and JTK Amplitude Q1 respectively. *q*-values are the results of multiple testing corrections of *p*-values using the Benjamini-Hochberg method. * *q*-value ≤0.1.

Examining heart rate, patients with a reverse dipping phenotype showed a 22% higher risk for developing delirium during their ICU admission (Odds Ratio: 1.22, 95% CI: 0.99-1.49, *q*=0.238, Figure 1 right) but that effect was not statistically significant.

### Dampened rhythms and reverse blood pressure dipping linked to increased risk of death

The mortality rate among patients with sufficient blood pressure recordings but excluding repeat ICU admissions was 15.9% (5,403 deaths within a month from ICU admission) compared to 16.3% (5,708 deaths) for the patient cohort meeting inclusion criteria for heart rate. Patients with a reverse dipping blood pressure profile had a 13% higher risk to die in the ICU setting within a month (Odds Ratio: 1.13, 95% CI: 1.02-1.26, *q*=0.07, Figure 2, Table S3). Also, compared to patients with a blood pressure amplitude in the first quartile, those in the third quartile were at a significantly lower risk of death (Odds Ratio 0.89 95% CI: 0.81-0.97, *q=*0.07) and there was a non-significant trend in the same direction for those in the second and fourth quartile. Furthermore, patients with a more robust heart rate rhythm belonging to the third amplitude quartile were protected from dying (Odds Ratio: 0.83, 95% CI: 0.75-0.92, *q*=0.004) as compared to those in the lowest heart rate amplitude quartile (Figure 2, Table S3), and patients in the second and fourth quartile had a non-significant trend in the same direction.

**Figure 2.**
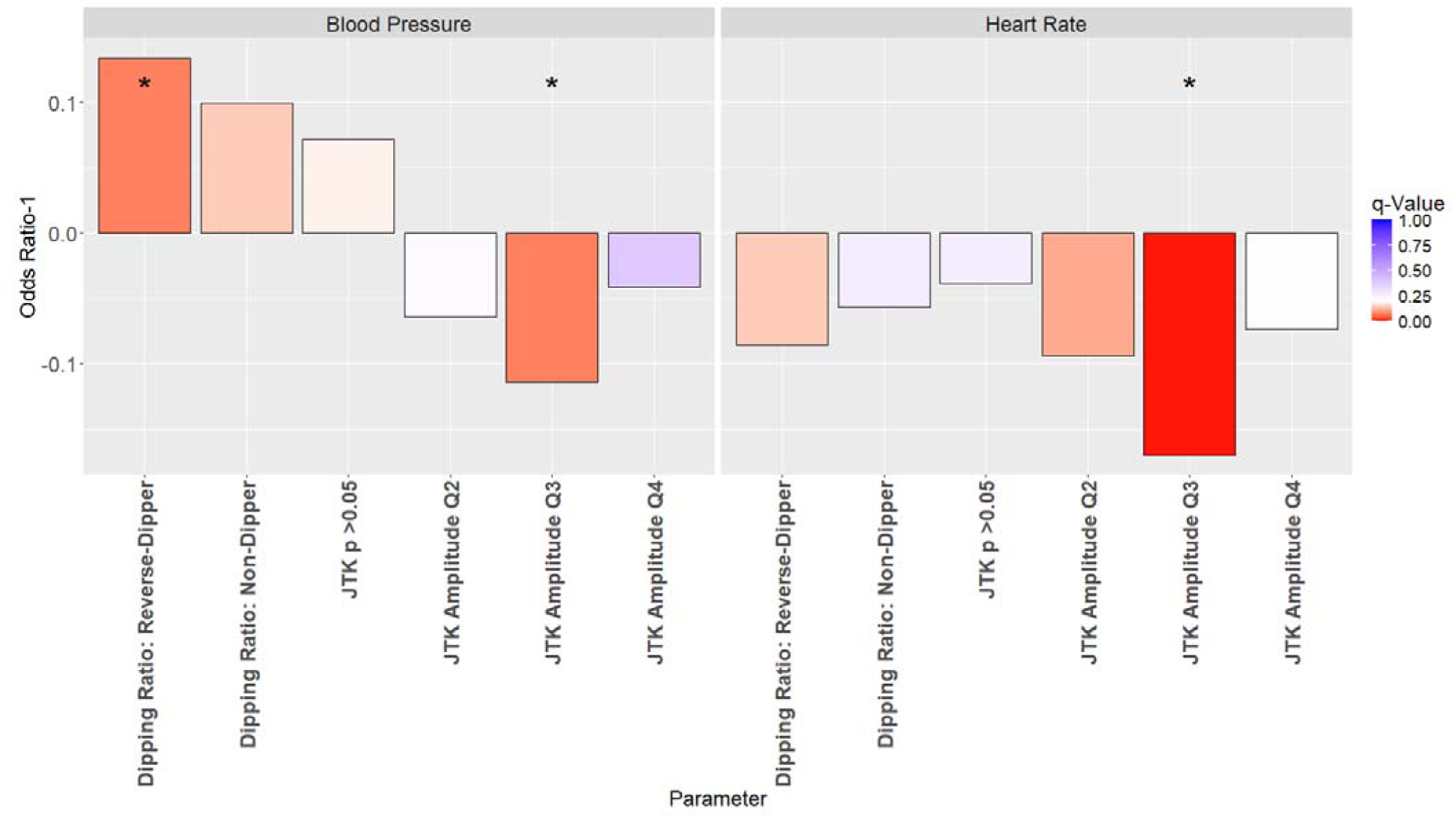
Associations between different rhythm parameters and 1 year mortality. The left panel displays results from the logistic regression models testing blood pressure rhythmic parameters, the right panel displays results from heart rate rhythmic parameters. Reference categories are Dippers, JTK *p-*value ≤0.05, and JTK Amplitude Q1 respectively. q-values are the results of multiple testing corrections of p-values using the Benjamini-Hochberg method. * q-Value ≤0.1.

### Reverse blood pressure dipping is associated with multi-system rhythmic disruptions

We explored whether reverse blood pressure dipping correlates with altered diurnal variability in biochemical processes assessed by clinical laboratory measurements (Figure S5). Because a single patient would rarely have enough repeated laboratory measurements to cover a 24-hour time series, we analyzed measurements across all patients belonging to either blood pressure dippers (controls) or reverse dippers (cases) in a mixed effects cosinor model. Among 101 laboratory tests recorded in the first 24 hours in patients with representative coverage of both day and night (at least 14 daytime and 6 nighttime measurements as used for blood pressure rhythm quality control criteria), a total of 45 laboratory tests demonstrated a statistically significant difference in rhythmicity in the cosinor fits comparing BP reverse dippers to dippers (*q*-value<0.01, Figure 3, Table S4, Figure S5). Candidates include hematocrit (*q*-value≤0.001), hemoglobin (*q*-value<0.001), arterial O2 pressure (*q*-value<0.001), platelet count (*q*-value<0.001), glucose (*q*-value<0.001), creatinine (*q*-value<0.001), prothrombin time (*q*-value<0.001), INR (*q*-value<0.001), lymphocytes (*q*-value=0.002), WBC (*q*-value≤0.001), PTT (*q*-value<0.001), and monocytes (*q*-value<0.001), all reported to be oscillatory though some with conflicting results particularly in PT, PTT, and INR.(24–28)

**Figure 3.**
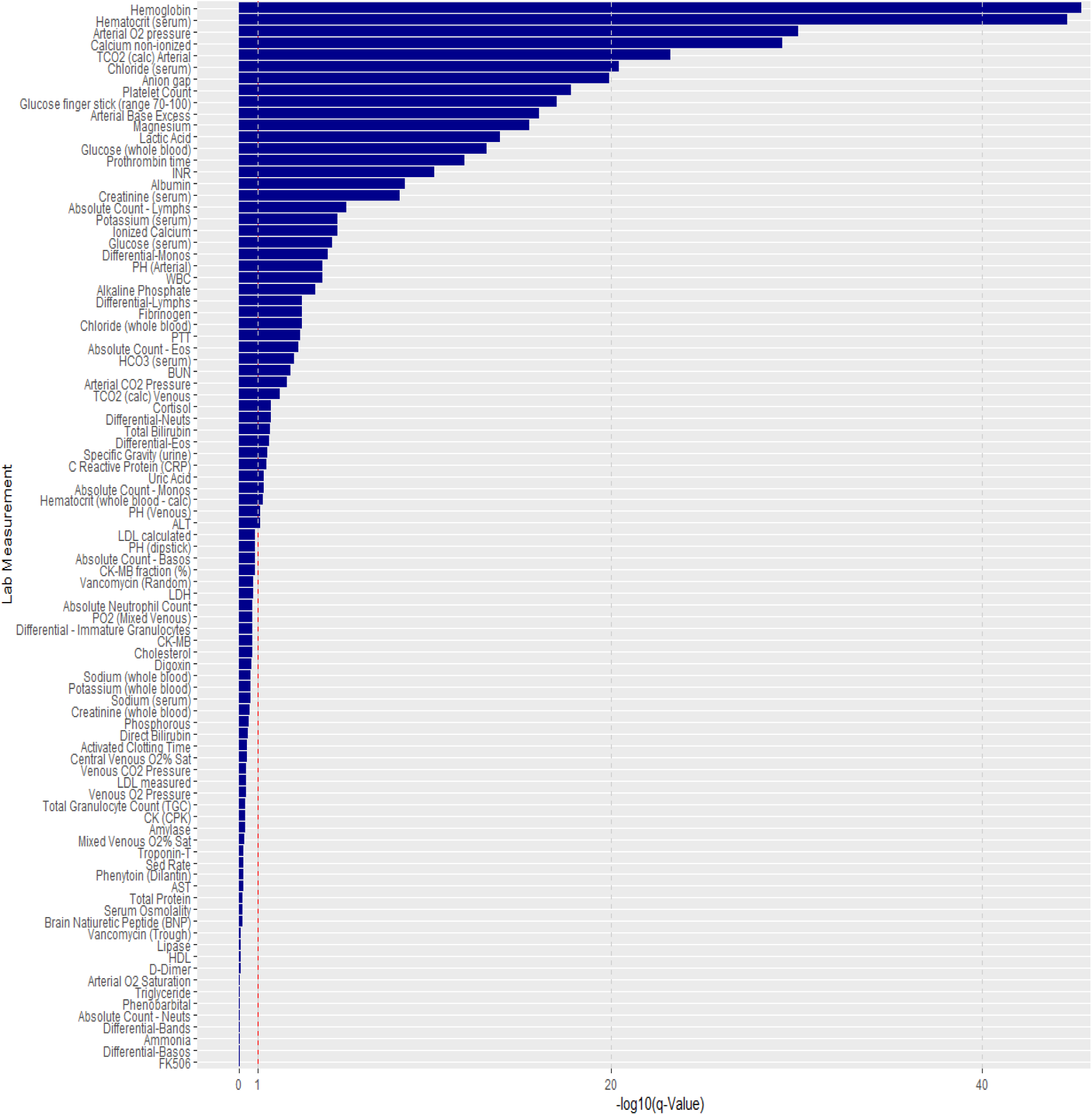
Significance levels of differences between blood pressure dipping statuses in cosinor fits of clinical laboratory measurements across the first 24 hours. The red line indicates a q-value cutoff of 0.1 for statistical significance. The actual values behind the graph are in Table S4.

Next, we examined rhythmic characteristics of these 45 labs to ascertain differences and commonalities between blood pressure dippers and reverse dippers (Figure 4A). Most of these clinical labs (n=29, 69%) displayed significant diurnal variability consistently in both groups (cosinor *q*-value≤0.1). About 14% (n=6) were only rhythmic in dippers. Among these were albumin and cortisol (Figure 4 B). The diurnal fluctuations in serum albumin levels in a healthy population likely contribute to time-specific blood pressure regulation through associated changes in oncotic pressure.(29) This functional relationship might deteriorate in reverse dippers as suggested by the lack of oscillatory albumin serum concentrations (Figure 4B). Cortisol’s diurnal secretion patterns in healthy volunteers likely drive the time-specific sensitization of the vasculature to the noradrenergic system (30, 31) a mechanism linked to blood pressure control.(32) Here, the lack of significant oscillations in reverse dippers compared to dippers may suggest that disrupted diurnal cortisol levels contribute to this pathology (Figure 4 B).

**Figure 4.**
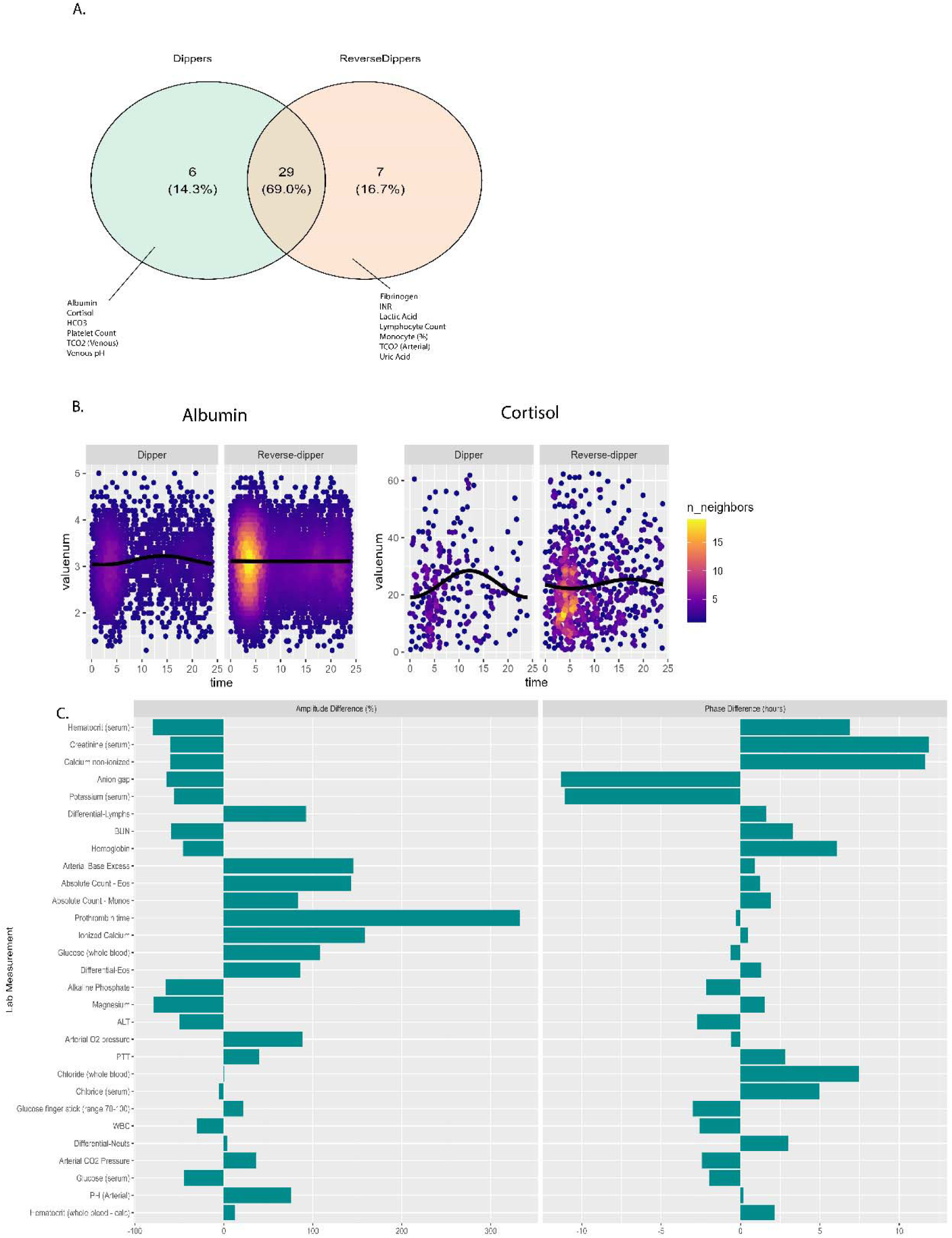
Amplitude and phase differences calculated from cosinor fits for laboratory measurements that showed statistically significant differences in cosinor fits between blood pressure dippers and reverse dippers. Values shown are relative to dippers and sorted by decreasing amplitude and phase differences. The actual values behind the graphs are in Table S4.

About 17% (n=7) lab tests showed significant oscillations only in patients with the reverse blood pressure dipping phenotype (Figure 4A). This is an unexpected finding and warrants further examination in future studies since time-of-day specific fluctuations have been reported in the healthy for the coagulation system,(33) immune system,(34) lactic acid and uric acid (35, 36). Conceivable, however, is that gain in diurnal fluctuations in the coagulation system are indicative of transient states of hypercoagulability, a known hemostatic abnormality in critical illness leading to poorer outcomes.(4, 37)

We then turned our focus to laboratory features significantly oscillating in both groups to assess differences in rhythmic components between dippers and reverse dippers. To examine the rhythmic components of the 29 clinical labs that were rhythmic in both dippers and reverse dippers we quantified cosinor amplitudes and acrophases (time of day of the peak) (Figure 4C, Table S4). Kidney function as measured by serum creatinine showed a 60% decrease in amplitude in reverse dippers compared to dippers. This was accompanied by a phase advance of 11.8 hours in reverse dippers compared to dippers. BUN similarly demonstrated a reduction in amplitude (60% in reverse dippers compared to dippers). Here, the phase differences were smaller (3.3 hours in dippers respectively compared to dippers). Lab values indicative of acid base balance showed higher amplitude fluctuations in reverse dippers compared to dippers, for instance, 145% higher amplitude in arterial base excess, 76% in arterial pH amplitude and 36% in arterial CO2 pressure amplitude. Electrolyte levels diverged, for example, by a 11-hour phase advance in non-ionized calcium levels in reverse dippers compared to dippers accompanied by a 60% amplitude dampening. This contrasted with a 159% increase in ionized calcium amplitude in reverse dippers with minimal phase changes. Several hemostatic parameters showed distinct rhythmic components in reverse dippers compared to dippers. Prothrombin time (PT) fluctuated with a 333% higher amplitude in reverse dippers and amplitude was 40% higher in partial thromboplastin time (PTT). Thus, reverse blood pressure dipping was associated on a population level with perturbed rhythms in clinical parameters of renal, metabolic, and hemostatic function.

## Discussion

Our present study demonstrates that patients with a reverse blood pressure dipping phenotype had a 40% greater risk of developing delirium during their ICU admission compared to patients with the blood pressure dipping phenotype. We showed that patients with reverse blood pressure dipping and dampened amplitudes for blood pressure and heart rate within the first 24 hours of ICU admission had a 13% higher risk of death within one month of their ICU admission. We postulate that the link between reverse BP dipping and delirium/death is accompanied by perturbed rhythms in clinical parameters of renal, metabolic, and hemostatic function, thus offering potential biomarkers for further interrogation of mechanism and risk prognosis.

Our finding that reverse blood pressure dipping precedes a diagnosis of delirium is supported by published data. We also take studies on long term cognitive decline and dementia into consideration since delirium is an established risk factor for developing chronic forms of cognitive impairment.(5) A longitudinal cohort study in older men in Sweden found a 64% higher risk for future dementia (and Alzheimer’s disease) among reverse dippers (Hazard Ratio: 1.64, 95% CI: 1.14-2.34).(38) The authors suggested reverse dipping as an independent risk factor for dementia (and Alzheimer’s).(38) Several cross sectional studies support the link between reverse blood pressure dipping and chronic forms of cognitive decline.(39–41) A preliminary report of a functional study using the head-up tilt test detected a poorer autonomic function in participants with delirium compared to controls.(42)

The higher mortality we observed in patients with reverse blood pressure dipping and blunted diurnal rhythms of blood pressure and heart rate highlights how time series of vital sign measurements routinely assessed in the critical care environment harbor information to understand patients’ risk profile. Though our results strengthen comparable findings in the eICU Collaborative Research Database (43), other studies in the same database surprisingly reported that a higher amplitude of heart rate oscillations was associated with elevated in-hospital mortality. (44)These discrepant findings might be in part driven by varying numbers of day– and nighttime measurements. Our study addresses this potential limitation by ensuring both nighttime and daytime periods were adequately represented in every included time series (at least 14 daytime and 6 nighttime measurements). Here, we advocate for a concerted effort to address research challenges in this space.(45)

Reverse blood pressure dipping is an alteration of the normal diurnal phenotype and is likely accompanied by time-specific changes in biochemical processes. Indeed, our results showed that reverse blood pressure dipping was associated with altered diurnal phenotypes in various clinical laboratory measurements. An important finding, for example, is the absence of rhythmic cortisol levels in reverse dippers (Figure 4). While we cannot directly assess catecholamines in this database, activation of the hypothalamic pituitary axis usually accompanies increased sympathetic tone.(4) Based on that we speculate that a disturbance in cortisol rhythms is also mirrored by a disturbance in sympathetic tone thatis oscillatory and plays a substantial role in regulating the diurnal variability in blood pressure.(46) Both elevated cortisol and sympathetic tone are associated with post-operative delirium.(4, 47) Reverse blood pressure dipping can thus be a consequence of disturbed autonomic nervous system rhythms that may increase the risk for developing delirium and other adverse outcomes. Given the specific roles the autonomic nervous system exerts on different physiologic systems including metabolism, renal function, and lung function (48), we speculate that the autonomic nervous system may thus represent a unifying candidate biomarker justifying further interrogation in future experimental studies. (10)

Our study has several strengths. We used a well curated, well documented, and commonly used database containing patient trajectories from a respected academic center. The size of the database permitted our application of strict inclusion criteria to ensure an adequate coverage of the first 24 hours for our rhythmic assessments. The database also recorded time-stamped CAM assessments, a well validated commonly used instrument to detect delirium. This enabled us to assign delirium diagnoses with a higher level of certainty than relying on diagnostic codes, which might have missed true positives.(4) Due to a low frequency of body temperature measurements and the consequent lack of confidence in their rhythmic assessments we had to abandon this biomarker. We also found that body temperature data were skewed; patients that had their temperature measured frequently were sicker. Another limitation inherent to this database is the observational design with a high degree of variability in patients, disease conditions, co-morbidities, and treatments. Overall, sicker patients might be over-represented since they likely had more interactions documented in the database due to higher care intensity. While pooling lab results from different patients is valuable to enable time-series statistics, options should be explored for repeat within-person biosampling. Lastly, the biomarker discovery is predetermined by the clinical lab panel.

## Conclusion

Reverse blood pressure dipping can be an early sign for the future development of delirium in the ICU and is accompanied by disrupted biorhythms across multiple organ systems. Dampened and reversed heart rate and blood pressure rhythms are associated with a higher risk for death in ICU patients. Considering the inclusion of these risk factors in preventive care may improve patient outcomes and reduce burden on the health care system.

## Availability of data and materials

Data used in this study is available for credentialed users at https://physionet.org/content/mimiciv/2.2/. All statistics derived from the dataset are available with the supplemental material attached to this manuscript.

The code developed for this study is freely available at: https://zenodo.org/records/14750969?preview=1&token=eyJhbGciOiJIUzUxMiJ9.eyJpZCI6ImEwMDZiNzRmLWE1NTAtNDVmZi1iMjczLTcwZDhjYmRhN2ZlOSIsImRhdGEiOnt9LCJyYW5kb20iOiJhZGRlMGQ3NjBiNWQ2MTM4YjZmOTE0MzNkZGQyOWYzZiJ9.2n0wyJLE_wMBQEiT5WR_NghvxgL5nakSEXJdGkw-VB6UCmantTQqiADE23S6GicaDAir2qfs-ST0Jl4IyAKw2g

## Ethics declarations

The MIMIC IV study was approved by the institutional review board at the Beth Israel Deaconess Medical Center and was granted a waiver of informed consent.

## Consent for publication

Not applicable

## Author Contributions

Conceptualization: NE, CS. Methodology: NE, CS, TB, AM, MG. Supervision: GF, CS. Writing original draft: NE, CS. Writing-review and editing: CS, GF, TB, AM, MG. All authors read and approved the final manuscript.

## Abbreviations

BP: Blood Pressure
CAM: Confusion Assessment Method
JTK: Jonckheere-Terpstra-Kendall
HR: Heart Rate
OASIS: Oxford Acute Severity of Illness Score
PT: Prothrombin Time
PTT: Partial Thromboplastin Time

## Data Availability

Data used in this study is available for credentialed users at https://physionet.org/content/mimiciv/2.2/. All statistics derived from the dataset are available in with the supplemental material attached to this manuscript.
The code developed for this study is freely available at: https://zenodo.org/records/14750969?preview=1&token=eyJhbGciOiJIUzUxMiJ9.eyJpZCI6ImEwMDZiNzRmLWE1NTAtNDVmZi1iMjczLTcwZDhjYmRhN2ZlOSIsImRhdGEiOnt9LCJyYW5kb20iOiJhZGRlMGQ3NjBiNWQ2MTM4YjZmOTE0MzNkZGQyOWYzZiJ9.2n0wyJLE_wMBQEiT5WR_NghvxgL5nakSEXJdGkw-VB6UCmantTQqiADE23S6GicaDAir2qfs-ST0Jl4IyAKw2g

## Supplementary Figures

**Figure S1.**
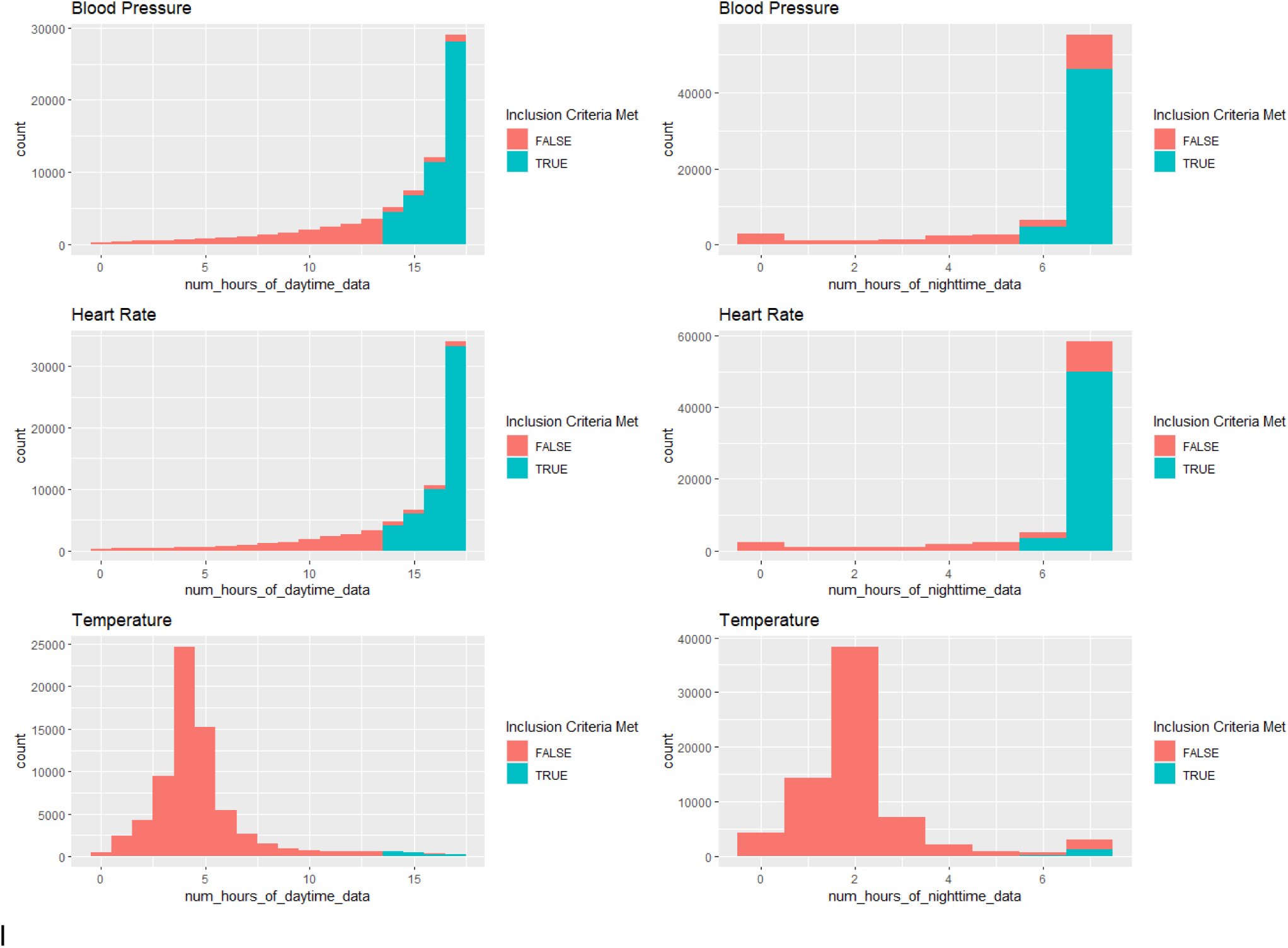
Histograms showing the number of admissions having each maximum number of hours of data recording in the daytime (left) and the nighttime (right). The blue bars represent the admissions that fulfilled our time series inclusion criteria, and the orange bars represent the admissions that do not meet the criteria. The majority of admissions have blood pressure and heart rate time series data that fulfil the inclusion criteria while only a minority of cases meet the criteria in temperature. Note the different scales of x and y axes.

**Figure S2.**
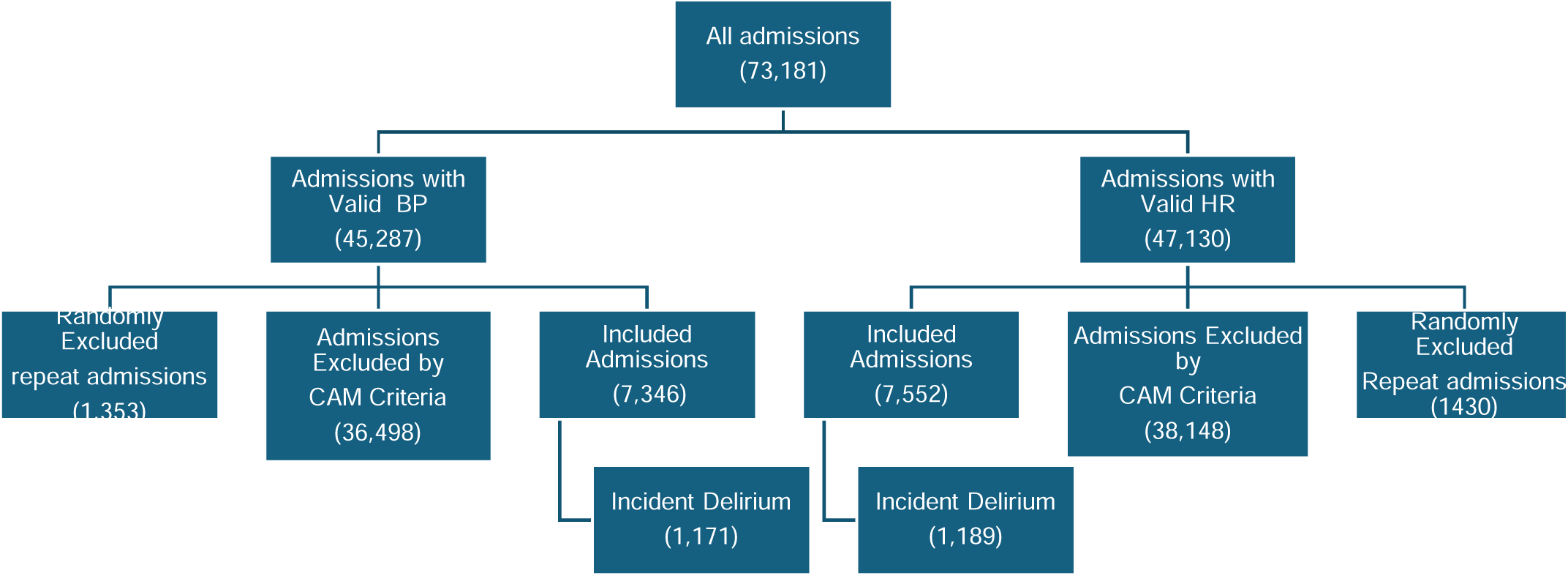
Flow chart representing the selection of admissions included in the delirium logistic regression analysis for each vital sign. Admissions included in the delirium analysis were those with a negative delirium assessment by CAM in the first 24 hours and a valid repeat CAM after that. Patients with multiple admissions fulfilling these criteria had one admission chosen at random.

**Figure S3:**
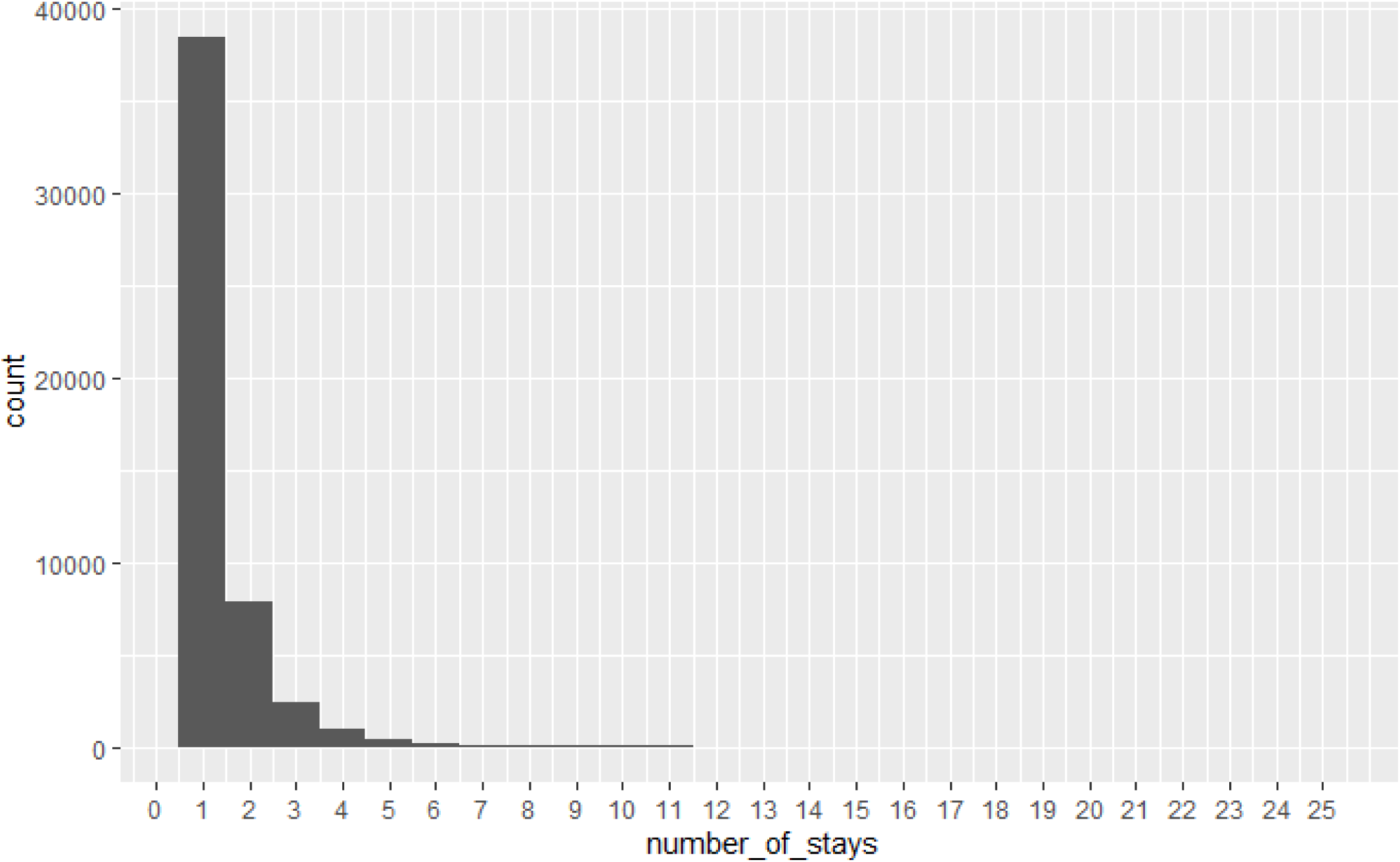
Histogram showing the distribution of patients across the number of admissions per patient.

**Figure S4.**
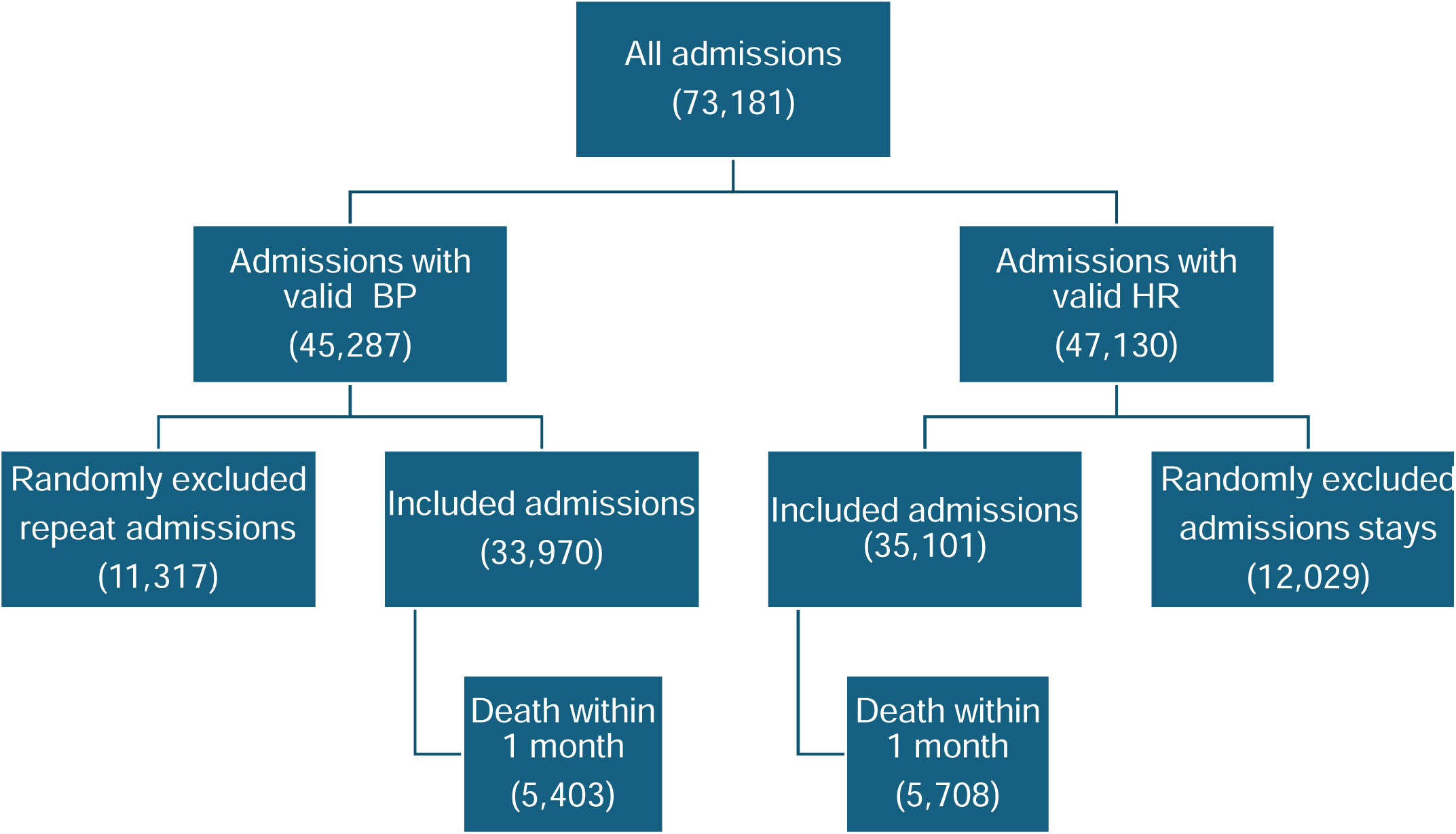
Flow chart representing the selection of admissions included in the 1-month mortality logistic regression analysis for each vital sign. Admissions included in the mortality analysis were those fulfilling our rhythm quality control criteria. Patients with multiple admissions fulfilling these criteria had one admission chosen at random.

**Figure S5:**
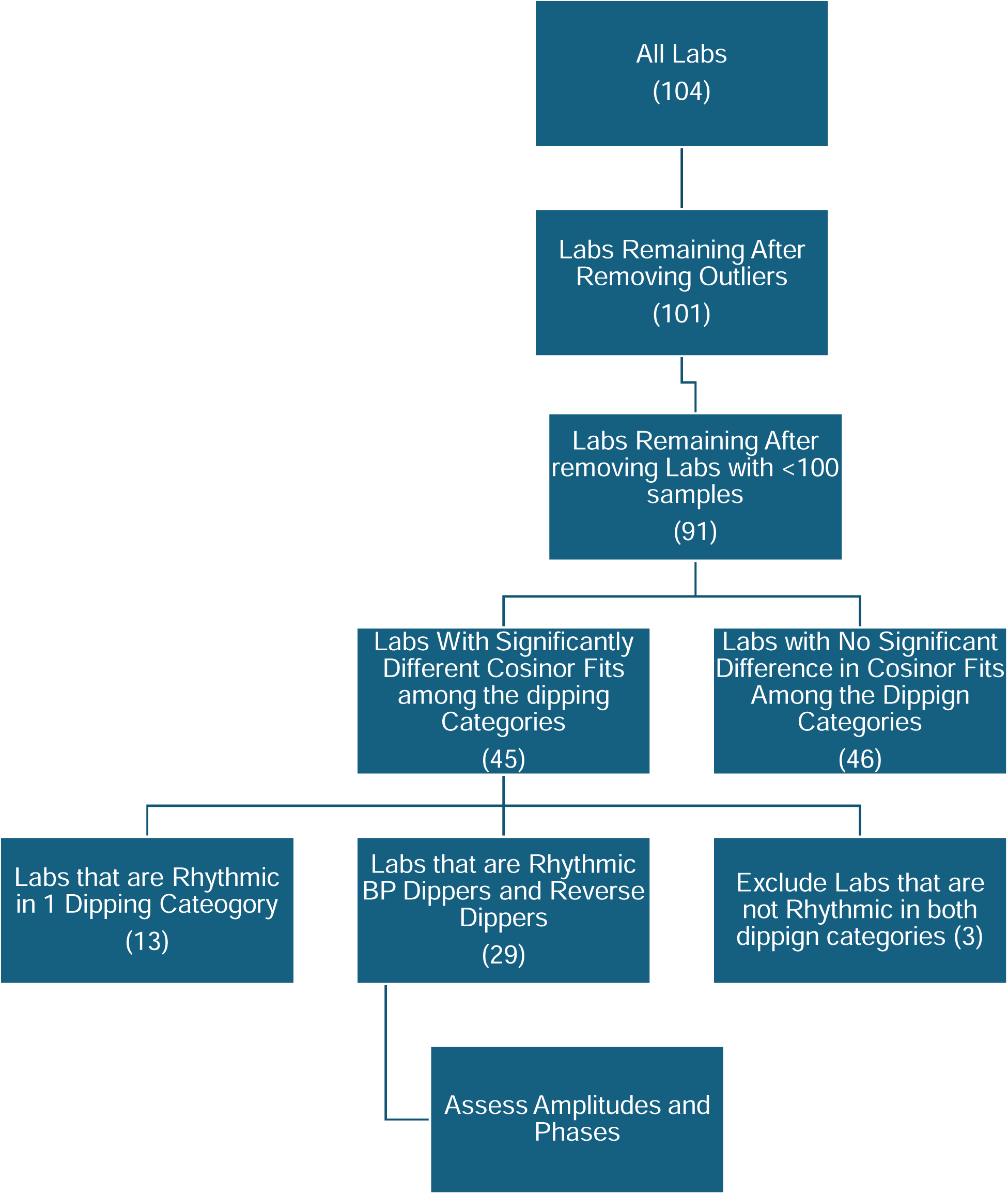
Flow chart representing the analysis process of the clinical lab dataset. The corresponding results are shown in figures 3 and 4.

## Supplementary Tables

**Table S1.** Amplitude quartiles and their corresponding minima and maxima.

| Vital Sign | Amplitude<br>Quartile | Minimum<br>Amplitude | Maximum<br>Amplitude | Mean<br>Amplitude | Amplitude<br>SD |
| --- | --- | --- | --- | --- | --- |
| Blood<br>Pressure<br>(mmHg) | 1 | 0.00 | 5.90 | 4.53 | 1.01 |
|  | 2 | 5.91 | 8.40 | 7.10 | 0.64 |
|  | 3 | 8.41 | 11.30 | 9.57 | 0.81 |
|  | 4 | 11.31 | 48.79 | 15.36 | 4.26 |
| Heart Rate<br>(bpm) | 1 | 0.00 | 3.50 | 2.20 | 0.83 |
|  | 2 | 3.50 | 5.69 | 4.57 | 0.75 |
|  | 3 | 5.70 | 8.49 | 7.19 | 0.79 |
|  | 4 | 8.52 | 49.67 | 12.89 | 4.50 |

**Table S2:**
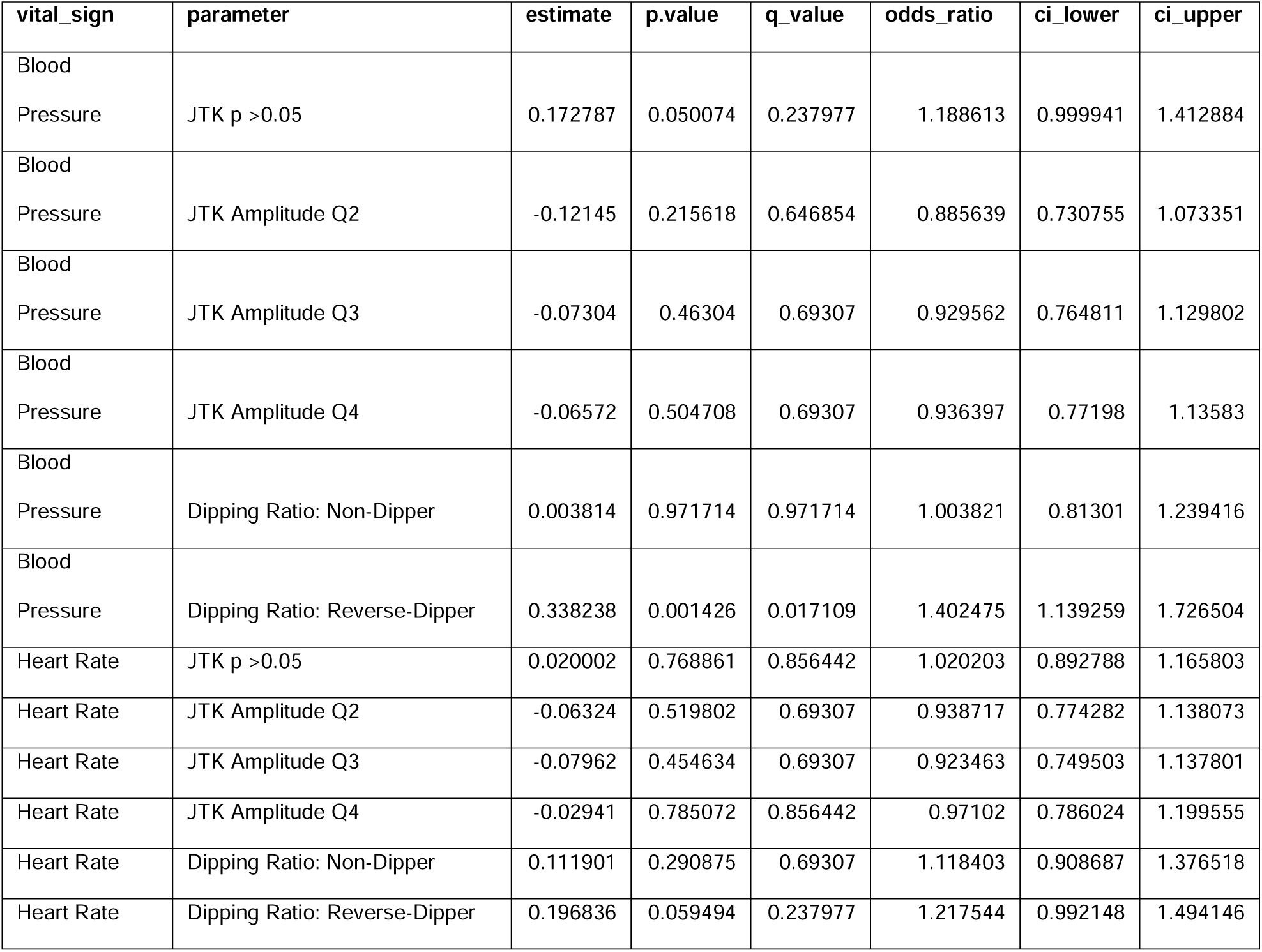
Numeric results of the delirium logistic regression models represented graphically in figure 1.

| vital_sign | parameter | estimate | p.value | q_value | odds_ratio | ci_lower | ci_upper |
| --- | --- | --- | --- | --- | --- | --- | --- |
| Blood Pressure | JTK p >0.05 | 0.172787 | 0.050074 | 0.237977 | 1.188613 | 0.999941 | 1.412884 |
| Blood Pressure | JTK Amplitude Q2 | -0.12145 | 0.215618 | 0.646854 | 0.885639 | 0.730755 | 1.073351 |
| Blood Pressure | JTK Amplitude Q3 | -0.07304 | 0.46304 | 0.69307 | 0.929562 | 0.764811 | 1.129802 |
| Blood Pressure | JTK Amplitude Q4 | -0.06572 | 0.504708 | 0.69307 | 0.936397 | 0.77198 | 1.13583 |
| Blood Pressure | Dipping Ratio: Non-Dipper | 0.003814 | 0.971714 | 0.971714 | 1.003821 | 0.81301 | 1.239416 |
| Blood Pressure | Dipping Ratio: Reverse-Dipper | 0.338238 | 0.001426 | 0.017109 | 1.402475 | 1.139259 | 1.726504 |
| Heart Rate | JTK p >0.05 | 0.020002 | 0.768861 | 0.856442 | 1.020203 | 0.892788 | 1.165803 |
| Heart Rate | JTK Amplitude Q2 | -0.06324 | 0.519802 | 0.69307 | 0.938717 | 0.774282 | 1.138073 |
| Heart Rate | JTK Amplitude Q3 | -0.07962 | 0.454634 | 0.69307 | 0.923463 | 0.749503 | 1.137801 |
| Heart Rate | JTK Amplitude Q4 | -0.02941 | 0.785072 | 0.856442 | 0.97102 | 0.786024 | 1.199555 |
| Heart Rate | Dipping Ratio: Non-Dipper | 0.111901 | 0.290875 | 0.69307 | 1.118403 | 0.908687 | 1.376518 |
| Heart Rate | Dipping Ratio: Reverse-Dipper | 0.196836 | 0.059494 | 0.237977 | 1.217544 | 0.992148 | 1.494146 |

**Table S3:** Numeric results of the mortality logistic regression models represented graphically in Figure 2.

| vital_sign | parameter | estimate | p.value | q_value | odds_ratio | ci_lower | ci_upper |
| --- | --- | --- | --- | --- | --- | --- | --- |
| Blood Pressure | JTK p >0.05 | 0.068902 | 0.108148 | 0.185397 | 1.071332 | 0.984957 | 1.165281 |
| Blood Pressure | JTK Amplitude Q2 | -0.06583 | 0.163673 | 0.21823 | 0.936287 | 0.853444 | 1.027171 |
| Blood Pressure | JTK Amplitude Q3 | -0.12083 | 0.013475 | 0.071496 | 0.886185 | 0.805193 | 0.975324 |
| Blood Pressure | JTK Amplitude Q4 | -0.04166 | 0.387489 | 0.387489 | 0.959197 | 0.872719 | 1.054245 |
| Blood Pressure | Dipping Ratio: Non-Dipper | 0.094311 | 0.072843 | 0.145687 | 1.098901 | 0.991301 | 1.218181 |
| Blood Pressure | Dipping Ratio: Reverse-Dipper | 0.125297 | 0.017874 | 0.071496 | 1.133485 | 1.021833 | 1.257337 |
| Heart Rate | JTK p >0.05 | -0.03946 | 0.231283 | 0.263046 | 0.961306 | 0.901156 | 1.02547 |
| Heart Rate | JTK Amplitude Q2 | -0.09864 | 0.037281 | 0.111844 | 0.906067 | 0.825741 | 0.994207 |
| Heart Rate | JTK Amplitude Q3 | -0.18568 | 0.000312 | 0.003748 | 0.830543 | 0.75079 | 0.918769 |
| Heart Rate | JTK Amplitude Q4 | -0.07606 | 0.134975 | 0.202462 | 0.926765 | 0.8388 | 1.023954 |
| Heart Rate | Dipping Ratio: Non-Dipper | -0.05802 | 0.241125 | 0.263046 | 0.943629 | 0.856379 | 1.039767 |
| Heart Rate | Dipping Ratio: Reverse-Dipper | -0.08966 | 0.068343 | 0.145687 | 0.914239 | 0.830209 | 1.006775 |

**Table S4:** External csv file with all the statistics from the cosinor fits and rhythm difference tests used to graph figures 3 and 4.

